# Systemic and structural barriers to effective infection prevention and control practices in a teaching and referral hospital in Kenya: A qualitative study

**DOI:** 10.64898/2026.07.22.26358730

**Authors:** Sakhile V. Ndwandwe, Abigail W. Nyanjui, Langelihle Ndebele, Racheal Kimani, Janet Ngethe, Nicole Wamaitha, Cynthia Nduta, Gladys G Njoroge, Samuel M. Mbugua, Jesse Gitaka

**Affiliations:** School of Pharmacy and Health Sciences, United States International University-Africa, Nairobi, Kenya; Ministry of Health, Kiambu County Government, Kenya.; Mama Ngina University College, Kenya; Mount Kenya University, Kenya

## Abstract

**Background:** Healthcare-associated infections (HAIs) are a leading adverse event in healthcare delivery. Despite Kenya’s robust National Infection Prevention and Control (IPC) Policy and Strategic Plan, a profound policy-to-practice gap persists at the institutional level. This study explored context-specific structural, behavioral, and administrative barriers hindering IPC implementation at a public referral hospital to inform localized policy tailoring.

**Methods:** We conducted a cross-sectional qualitative study at a level V teaching and referral hospital in Kenya. Seven key informant interviews were held with five IPC committee members/providers and two departmental managers. Data from audio recordings and field notes were transcribed, translated, and analyzed via thematic content analysis using QSR NVivo V.12.

**Results:** Seven major themes emerged: (1) IPC Training and Knowledge Retention, (2) Workforce Strain and Triaging of IPC, (3) Resource Scarcity and a Disabling Environment, (4) The Community-Hospital Interface, (5) Governance and Policy-to-Practice Lag, (6) IPC Surveillance and Monitoring, and (7) Behavioral Drivers (Attitude, Ignorance, Accountability). Financial and human resource constraints directly undermine continuous medical education, workforce ratios, WASH infrastructure functionality, and the local adaptation of national guidelines.

**Conclusion:** A substantial policy-to-practice gap persists at the facility level due to severe structural, resource, and behavioral constraints. To bridge this gap, hospital leadership must transition from reactive crisis management to proactive, continuous medical education and sustainable WASH infrastructure development. Effective IPC implementation requires national frameworks to be dynamically adapted and tailored to the operational realities and resource limitations faced by frontline healthcare workers.

## INTRODUCTION

Healthcare-associated infections (HAIs) remain a daily threat in every hospital and clinical setting, affecting patients, causing immense suffering, driving higher healthcare costs, and hindering efforts to achieve high-quality health [1,2]. According to the World Health Organization antibiotic-resistant infection cases were estimated at 136 million annually, about 119 million of these occurred in middle-income countries, and patients with healthcare-associated sepsis were estimated to face a 24.4% mortality [2,3]. In sub-Saharan Africa, HAIs are often an underestimated public health burden with an estimated pooled prevalence of 12.9%–13%, which is significantly higher than that of developing nations [4]. In Kenya, healthcare-associated infections (HAIs) represent a significant but often under-reported public health burden. Data shows that about 10%– 25% of hospital admissions in government health facilities are affected by HAIs [5].

Infection Prevention and Control (IPC) is a scientific, evidence-based approach and practical solution designed to prevent harm caused by infections to patients, health workers, and visitors in health facilities and communities [6]. During the Seventy- seventh World Health Assembly, a global action plan and monitoring framework for Infection Prevention and Control (IPC) for 2024–2030 was approved, providing clear actions, indicators, and targets to support member states in improving national and facility-level Infection Prevention and Control (IPC) actions [2]. The past decade has also seen significant investments in IPC activities across sub-Saharan Africa, including the development of global and national guidelines [3]. The COVID-19 pandemic led to significant changes in water, sanitation, and hygiene (WASH) policies and infrastructure [7,8]. However, evidence shows serious issues with adherence to standard IPC guidelines and the urgent need to reform IPC practices in sub-Saharan Africa [9].

In December 2010, the Kenyan Ministry of Public Health and Sanitation, along with the Ministry of Medical Services, published the national infection prevention and control guidelines for health and care services [10]. This proposed a comprehensive framework to standardize practices across all health sectors with guidelines focusing on key areas such as standard precautions, additional (transmission-based) precautions, environmental and operational management, specialized care and clinical areas, and management and occupational health [10,11]. The World Health Organization (WHO) [12] issued guidelines for IPC at the national and facility level, outlining eight core components for the implementation of effective IPC across all countries and facilities, and estimated that adherence to these IPC practices can reduce healthcare-associated infections (HAIs) rates by 30%. However, because feasibility is highly context- dependent, guidelines must be dynamically informed by, and adapted to, the operational barriers faced in low-resource settings [13]. In the context of Infection Prevention and Control, structural barriers refer to physical or logistical flaws in infrastructure and resources (e.g., lack of handwashing sinks or isolation rooms), while systemic barriers refer to broader organizational, policy, or societal shortcomings (e.g., underfunding, supply chain breakdowns, or poor safety cultures) [14–16].

Kenya is currently positioned as a regional leader in the African context for IPC policy development, having successfully established a robust national framework through its 2021–2025 Strategic Plan and National IPC Policy [5]. According to the World Health Organization (WHO) Global Report, while Kenya demonstrates advanced progress in creating technical guidelines and formalizing a national IPC program, it still faces an implementation gap common in the Regional Office for Africa (AFRO) region, where policy at the top does not always translate to consistent execution at the facility level [17]. Key challenges remain in scaling subnational monitoring, addressing workforce shortages, and improving the built environment, specifically water, sanitation, and hygiene (WASH) infrastructure, as the country moves from a basic to an intermediate level of global IPC competency [17,18]. While these policies exist at the national level, there seems to be an implementation-policy gap at facility A [9,18]. A study by [19] found that the availability of IPC supplies served as a significant determinant of compliance with IPC guidelines at a local facility. In Kenya a level V hospital is one that provides a wider variety of surgical and medical specialties, multiple operating theaters, and 24-hour advanced diagnostics (such as computed tomography (CT) scans). It is affiliated with universities to train medical graduates, nurses, and paramedical staff. It serves as an internship center for newly qualified doctors and clinical officers. And, requires a minimum inpatient capacity of over 100 to 300 beds [20]. The qualitative research presented in this paper sought to explore the context-specific barriers to implementing effective IPC practices and identify barrier-specific factors hindering IPC practices at a level V facility, in Kenya, for policy adaptation and tailoring.

## METHODS

We employed a cross-sectional qualitative study design at facility A, aimed at gaining lucid insights into health system challenges in infection prevention and control. Key informant Interviews were conducted to allude to the perspectives of health providers on current IPC practices and explored barriers. A purposive sampling strategy was employed to select participants who would provide in-depth insights into systemic and structural barriers. Specifically, a total of seven participants were interviewed, five of whom belonged to the IPC committee, and two were department managers. This non- probability method was chosen to ensure that the sample possessed the necessary lived experiences to address our research objectives effectively.

## DATA COLLECTION

Interviews were conducted in English and Swahili and audiotaped using recorders. The researchers acknowledged their positionality as an influence on the research process and employed reflexivity practices, including an audit trail, consistent interview procedures, and iterative analysis, to minimize bias and ensure findings were grounded in participants’ perspectives. Before interviews began, participants were provided with background information on the purpose of the interviews as well as the promise of confidentiality. Participants were requested to sign a consent form before beginning each interview.

## ETHICAL CONSIDERATIONS

We received a National Commission for Science, Technology and Innovation (NACOSTI) research license, administrative approvals from the Kiambu County health department, as well as approval from the facility itself.

## DATA ANALYSIS

Qualitative data was translated, transcribed, and exported to QSR NVivo V.12 for analysis. 3 members of the research team used an iterative analysis process to develop a coding framework and later a thematic framework to classify and organize data into emergent themes.

## RESULTS

Participants described challenges to IPC that were explored along seven major themes: (1) IPC Training and Knowledge Retention, (2) Workforce Strain and the Triaging of IPC, (3) Resource Scarcity and the Disabling Environment, (4) The Community-Hospital Interface and Public Sensitization, (5) Governance Structure and the Policy-to-Practice Lag, (6) IPC Surveillance and Monitoring, (7) Behavioral Drivers: Attitude, Ignorance, and Professional Accountability. Participants also spoke of potential IPC solutions.

## IPC TRAINING AND KNOWLEDGE RETENTION

### 1. SUB-OPTIMAL FREQUENCY AND KNOWLEDGE DECAY

The participants’ reflection on training indicated that formal IPC training occurred sporadically, falling far short of recommended frequencies. Several respondents noted that their last comprehensive training took place over two years ago, leading to significant knowledge decay and the erosion of standard competencies.

> *“Yes, we were trained like two years ago on IPC… we need to be refreshed… so that people can acquire knowledge that does not dry up, as people are reminded every day about IPC protocols.”* — **IPC Committee Member**

This lack of regular reinforcement results in health providers struggling to recall specific procedures, potentially impacting patient safety at the point of care. One participant expressed the difficulty of maintaining vigilance without consistent reminders:

> *“When did I last receive training in IPC? I can’t even recall, but I know there was an IPC sensitization. But I can’t recall when.”* — **IPC team, Procurement**

### 2. REACTIVE TRAINING CULTURE AND THE KNOWLEDGE GAP IN PRE-SERVICE STUDENTS ON CLINICAL APPRENTICESHIP

The data suggests that the facility employs a reactive training model rather than a proactive risk reduction strategy. Sensitization is often triggered by the emergence of an outbreak or an observed crisis, rather than being part of a structured, preventative education program.

> *“We usually conduct sensitization… especially when we see a gap or any issues of infection… e.g., an outbreak, or when we anticipate one, we prepare ourselves.”* — **IPC Focal Person**

> *“Monitoring only happens when there is a spike in surgical site infections. We don’t track baseline HAIs continuously”*-**IPC Committee Member**
>
> Furthermore, participants identified a systemic gap in the pre-service education of rotating clinical students. Many students reportedly arrive at the facility with little to no foundational knowledge of IPC, placing an additional training burden on an already strained hospital staff.

> *“In many cases, you find that students come in and most of them do not know anything about IPC… they encounter some of these things here.”* — **IPC Committee Member**

This finding underscores an urgent need for IPC principles to be integrated into theoretical training within medical and nursing schools before students enter the clinical environment.

### 3. LOW PRIORITIZATION OF INDUCTION AND ONBOARDING PROCESSES

The data suggests that the time-to-competency for new staff has been severely shortened. What used to be a thorough induction has become a rushed formality, leading to immediate compliance issues from day one.

> *“…at times the induction may be done maybe one day, three days. Initially, it used to take a lot of days but nowadays the time is quite limited… so when it comes to limitations, there is no good compliance.”* — **IPC Team, Procurement**

This shows that poor IPC practice starts at the very beginning of a staff member’s tenure due to administrative time constraints.

### 4. EXCLUSION OF CASUAL AND NON-CLINICAL STAFF

A major gap identified is that IPC training is often reserved for qualified clinicians, leaving casual workers, who handle waste and cleaning, vulnerable and uninformed.

> *“Since I came in March, I have not had any training… but for the casuals they are not trained. We need to create awareness of the IPC protocols so they can take care of themselves.”* — **IPC Committee Member**

IPC is only as strong as its weakest link; excluding casual staff creates a significant blind spot in hospital safety.

### 5. STAGNANT GOVERNANCE AND LEADERSHIP GAPS

Training and sensitization require active leadership. The data indicates that a lack of committee meetings and departmental leadership oversight results in a frozen training environment where staff *“don’t know where they are”* in terms of current standards.

> *“We have not had a meeting since I reported here in March of this year, and this is something that needs to be supported.”* — **IPC Committee Member**

> *“For those who are there… the HODs (Heads of Department) should take up themselves to train their groups before they enter.”* — **Nurse Manager, Newborn Unit**

This emphasizes that IPC education should not just come from the top but needs to be a departmental responsibility led by departmental heads.

### 6. THE KNOWLEDGE DRY-UP AND DECENTRALIZED LEARNING

Participants described a phenomenon where IPC knowledge dries up if it is not constantly replenished. The solution proposed by the data is a move away from centralized one-off out of town workshops toward local, frequent forums.

> *“We also have to have as many CMEs as possible to create awareness closer to the people. We should bring in the aspect of many forums so that people can acquire knowledge that does not dry up, as people are reminded every day about IPC protocols in the facilities.”* — **IPC Committee Member**

> *“We had gone out of town sometime about two or three years ago… but since then we have not gone for another so I don’t know where we are now.”* — **IPC Committee Member**

This underscores the necessity of ongoing IPC training, as prolonged intervals between sessions inevitably cause healthcare workers’ theoretical knowledge to drift from their daily practice.

## WORKFORCE STRAIN AND THE TRIAGING OF IPC

### 1. STAFF-TO-PATIENT RATIOS AND THE EROSION OF ADHERENCE

Participants identified chronic understaffing as a fundamental barrier that fuels professional exhaustion and inconsistent IPC practices. High staff-patient ratios create a high-pressure environment where healthcare workers must constantly navigate competing clinical tasks, often at the expense of standardized infection protocols.

> *“And also, the numbers. Yes, the staff-patient ratios. At times, we are very constrained such that you may not be able to effectively adhere to the IPC guidelines.”* — **IPC Focal Person**

This shortage is particularly acute in specialized areas like the Newborn Unit, where the presence of only *one qualified nurse* forces a compromise in care quality. Furthermore, the lack of personnel limits the ability of staff to provide essential IPC education to caregivers and patients.

> *“One [barrier] is a shortage of health personnel to train these mothers and to train each and every one. Because the workforce is strained, sometimes you get one qualified nurse in this unit*.*”* — **Nurse Manager, Newborn Unit**

### 2. COMPETING TASKS AND SURVEILLANCE GAPS

The data highlights a significant conflict of duties for those tasked with oversight. Members of the IPC committee are rarely dedicated solely to infection control; instead, they manage full clinical workloads while attempting to conduct surveillance and planning. This dual burden prevents the committee from maintaining a consistent, around-the-clock 7. presence.

> *“People who are members of the committee have other tasks, so sometimes it’s hard for them to participate because they are alone in the ward.”* — **IPC Focal Person**

> *“we need to have an IPC committee that works around the clock, because currently it does not work around the clock due to issues with personnel.”*— **IPC committee member**

Currently, the facility operates with one IPC focal person for 500 beds, a 100% deviation from the national guideline of one coordinator per 250 beds. This deficit makes it operationally impossible to effectively assess daily practices or maintain continuous medical education (CME).

> *“The guidelines state that at least 250 beds should have 1 IPC focal person or coordinator; if it’s more than that, we should have 2. Here you find one focal person against 500 beds. This person is not just dealing with IPC alone but also their other tasks…”*— **IPC focal person**

### 3. STAFF SHORTAGE AND COMPROMISE IN IMPLEMENTATION OF IPC PROTOCOLS

In the context of severe understaffing, participants described a triaging mindset where infection prevention is bypassed in favor of immediate life-saving interventions. When emergencies arise, the time required for proper IPC measures, such as changing gloves or hand hygiene, is viewed as a luxury the staff cannot afford.

> *“Maybe you are in the middle of a procedure, and there is another patient who needs a lot of attention, there is an emergency so definitely… see, you remove gloves, sanitizer if it’s within easy reach you are able to sanitize… At times there may be no time.”-***IPC Committee Member**

This highlights that IPC is often viewed as a time-dependent luxury rather than a non- negotiable step in emergency care. In critical units like the Newborn Unit, the presence of only a single qualified staff member creates a total collapse of the buddy system or peer-supervision necessary for IPC.

> *“Because the workforce is strained, sometimes you get one qualified nurse in this unit.”-***IPC focal person**

When a nurse is alone in the ward, there is no one to assist with tasks, increasing the likelihood that they will skip hand hygiene or glove changes to move between patients quickly.

### 4. EDUCATION DISPLACEMENT

The data shows that workforce strain does not just affect clinical actions but also stops the transmission of knowledge to patients (caregivers/mothers).

> *“The shortage of health personnel to train these mothers and to train each and everyone.”*—**Nurse manager, Newborn Unit**

This identifies a hidden outcome of understaffing: the inability to engage in Patient and Family Education, which is a core component of the WHO IPC standards.

## RESOURCE SCARCITY AND THE DISABLING ENVIRONMENT

### 1. INFRASTRUCTURE DECAY AND PROCUREMENT LATENCY

Participants identified resource constraints as the most pervasive challenge, describing an environment that fails to enable standard IPC practices. A primary driver of this scarcity is procurement latency and delayed maintenance, which leaves critical infrastructure, such as handwashing stations, non-functional for extended periods.

> *“For example, if there is a leaking handwashing basin and it takes a lot of time to be repaired… the female surgical [ward], all the 6 sinks are broken. It’s a challenge when it comes to IPC.”* — **IPC Focal Person**

> *“In the female surgical ward, all 6 sinks are completely broken. At times we have to go to the kitchen to wash hands… there are no paper towels, so after washing, you wipe your hands on your clothes or uniform…”* — **Nurse Manager, Newborn Unit**.

This systemic failure often forces healthcare workers to seek improvised solutions, such as using kitchen sinks for clinical hand hygiene, which compromises the aseptic chain.

### 2. CRISIS-DRIVEN SUPPLY CHAINS AND PPE SHORTAGES

The data suggests that the provision of Personal Protective Equipment (PPE) is often crisis-driven rather than consistent. Participants noted that while specialized equipment like eye shields were available during the COVID-19 pandemic, these supplies have since been discontinued, even in high-risk areas like the theater and labor wards.

> *“Sometimes we have a shortage of PPEs. Like now in a place like let’s say labor ward and theatre. You’re supposed to have the goggles to protect your eyes or that shield, there’s a shield that was provided during COVID era but those PPEs are no longer being provided”* — **IPC Committee Member**

Furthermore, the quality of available supplies, specifically proper soap and paper towels, is often inconsistent, directly undermining hand hygiene, which is globally recognized as the most critical IPC practice [8].

> *“…lack of IPC materials like, you can have soap and water but there are no paper towels. We have the sanitizers which are not enough…”*—**Nurse manager, New Born Unit**

### 3. IMPROVISED PRACTICES AND SAFETY COMPROMISES

In the absence of adequate resources, staff are forced to adopt workarounds that increase the risk of cross-contamination. The lack of drying materials, for instance, leads staff to dry their hands on their own clinical attire.

> *“Then on the same note we do not have hand towels to dry our hands so you get that someone is trying to wash their hands but they are trying to wipe the same hands in their clothes. And remember you are working and your clothes may have microorganisms so probably you’ll be doing zero work.”* — **IPC In-Charge**

Resource shortages also extend to waste management, where the lack of color-coded bin liners forces the dangerous practice of reusing liners, contradicting national biohazardous waste protocols.

> *“…the bin liners are not available the way they are supposed to be… sometimes we have to reuse these bin liners which is not very healthy.”* — **IPC In-Charge**

### 4. INADEQUATE POINT-OF-ENTRY BARRIERS

The data shows a failure to establish IPC boundaries at the entrances and exits of clinical departments. This potentially creates an environment where pathogens can move freely between wards and the public pointing to a structural failure in spatial IPC, where the hospital layout does not force hygiene compliance at critical transition points.

> *“We are supposed to have a hand-washing station at the entrance so that as they (healthcare personnel and the public) come in they perform hand hygiene… although that has not been done in most of the departments.”* — **IPC Committee Member**.

### 5. DECOUPLED SUPPLY AND INFRASTRUCTURE (THE HALF-MEASURE BARRIER)

The data reveals a decoupling of resources, where the facility may have the consumable (soap) but not the infrastructure (sink), or vice versa. This creates a half- measure environment where IPC is impossible to complete.

> *“In the female surgical ward, all 6 sinks are completely broken. At times we have to go to the kitchen to wash hands… there are no paper towels, so after washing, you wipe your hands on your clothes or uniform…”* — **Nurse Manager, Newborn Unit**.

> *“Go to the medical ward and there is only one hand washing station. It is not sufficient because of the population in that ward.”* — **IPC Focal Person**.

This emphasizes that IPC efficacy is determined by the weakest link in the infrastructure. One working sink for an entire ward population makes frequent hand hygiene geographically and logistically impossible.

## THE COMMUNITY-HOSPITAL INTERFACE AND PUBLIC SENSITIZATION

### 1. CAREGIVERS AS VECTORS AND VULNERABLE POPULATIONS

A recurring theme among participants was the recognition that the hospital environment is not a closed system. Health workers operate within a multitude that includes patients’ relatives and caregivers, who often lack the fundamental IPC knowledge required to navigate a clinical setting safely. Participants identified this population as a critical link in the chain of infection, capable of introducing community-acquired pathogens into the ward or carrying HAIs back to the community.

> *“The caregivers can bring infections from outside to the patients, and can carry infections outside. So, it’s a very critical population that needs to observe IPC practices when coming to the hospital…”* — **IPC Committee Member**

### 2. CHALLENGES IN MOVEMENT AND BEHAVIORAL CONTROL

The structural layout and management of the facility present a barrier to monitoring the public’s movement. Participants noted that once visitors enter the facility, there is a lack of oversight regarding their adherence to hand hygiene or their interaction with contaminated surfaces, which complicates the maintenance of an aseptic environment.

> *“When these people (patient relatives) come to see their relatives. No one controls how relatives move in the wards; they sit on beds and touch equipment…”* — **IPC Committee Member**

### 3. FORMALIZING CAREGIVER EDUCATION AND DISCHARGE PLANNING

To mitigate these risks, participants advocated for a structured approach to public sensitization. This includes creating dedicated forums to educate relatives on best practices during their visit and, crucially, how to continue infection prevention measures once the patient is discharged.

> *“We can create forums to educate the relatives. We can sensitize on IPC best practices… So, it’s a very critical population that needs to observe IPC practices.”* — **IPC Committee Member**

### 4. THE SOCIO-ECONOMIC AND PSYCHOLOGICAL BARRIERS OF VISITORS

The data reveals that visitors are not a uniform group; they arrive with varying educational backgrounds and are often in a state of crisis, which makes standard IPC messaging difficult to absorb.

> *“But we cannot say that they know what IPC is all about. They are from different backgrounds and are in a rush to get the help they need.”* — **IPC Committee Member**.

This highlights that ignorance among the public is often driven by the stress of the hospital visit and varying levels of health literacy.

### 5. SHIFT TOWARD FAMILY-CENTERED CARE IN SPECIALIZED UNITS

In units like the Newborn Unit, there is an emerging shift toward family-centered care, where mothers are not just visitors but essential partners in the patient’s recovery. This creates a functional necessity for IPC education to ensure safety during and after hospitalization.

> *“So that is their load because we cannot keep the baby here permanently. So, they also need to be educated… we are encouraging family-centered care, whereby we include the mothers.”* — **Nurse Manager, Newborn Unit**.

This positions IPC education as a tool for empowering caregivers, preparing them for the *“load”* of care post-discharge.

### 6. INEFFECTIVE TARGETING AND MISSED OPPORTUNITIES

Participants noted that current efforts are limited to basic hand hygiene reminders rather than a comprehensive approach to infection control, leaving the public under-utilized as partners in safety.

> *“Apart from maybe encouraging them to do hand hygiene… no. We have not been targeting them effectively.”* — **Member of IPC Committee**.

This identifies a missed opportunity where the hospital could be using the caregivers to help monitor the environment if they were properly sensitized.

## GOVERNANCE STRUCTURE AND THE POLICY-TO-PRACTICE LAG

### 1. TEMPORAL GAPS IN EVIDENCE-BASED UPDATES

Participants emphasized that IPC is an evolving scientific discipline requiring timely communication of evidence-based updates. However, a significant temporal gap exists between the development of national policies and their arrival at the facility level. This delay forces the hospital to operate under outdated protocols, which participants identified as a direct contributor to increased HAI rates. Participants also noted that while frameworks exist at national and county levels, “*laxity”* in the dissemination process prevents frontline workers from accessing current standards. In the absence of timely communication, healthcare workers rely on institutional memory and legacy methods.

> *“Everything is evolving and there is continuous research being done. So definitely, if people have the current and more updated information it’s very important to ensure you’re not **-**keeping the traditional practices.”* — **IPC Committee Member**

> *“These policies should come to us on time, because some of these policies were developed last year, and we are only receiving them this year. So, there is that laxity in how this information was disseminated for us to use in our work”* — **IPC Team, Procurement**

### 2. THE UNFUNDED MANDATE: POLICY VS. ENFORCEMENT POWER

A critical concern raised was the distinction between the *availability* of a policy and the *capacity* to implement it. Participants argued that guidelines without corresponding resource support are unenforceable. This highlights that effective governance requires not just the transmission of information, but the provision of the structural enablers, such as funding and supplies, needed to bring those policies to life.

> *“We need to be supported because even though we have these policies, we cannot enforce them.”* — **IPC Committee Member**

### 3. KNOWLEDGE OBSOLESCENCE AS A SAFETY RISK

There is an underlying theme that operating on old information is an active risk factor for HAIs. The need for CMEs to update is not just about professional development; it is seen as a protective measure for both patients and staff.

> *“There are also CMEs to update… it’s very important to ensure at least I mean, you’re not keeping the traditional practices.”* — **IPC Team, Procurement**

This frames the Policy Lag as a safety hazard. If the governance structure is slow, it inadvertently forces staff to practice sub-optimal medicine.

## IPC SURVEILLANCE AND MONITORING

### 1. REACTIVE VS. PROACTIVE SURVEILLANCE

A primary theme emerged regarding the need-basis nature of current monitoring efforts. Participants indicated that instead of a proactive, robust system, surveillance is often triggered only by specific incidents or suspected outbreaks. This is particularly evident in high-risk areas like Tuberculosis (TB) management:

> *“…especially the TB cases…I don’t know if they ever do surveillance. I think we do it on a need-basis. When you have issues now maybe is when we investigate it further.”* — **IPC Team Member**

### 2. BENCHMARKING AND ATTRIBUTION OF HAIS

The lack of continuous monitoring creates a significant gap in the facility’s ability to distinguish between community-acquired and healthcare-associated infections (HAIs).

Participants noted that without a consistent data baseline, it is nearly impossible to establish accountability or identify specific failure points in clinical procedures:

> *“Although sometimes it’s hard to tell that these catheter-associated urinary tract infections were gotten from us (the hospital), we have not been doing surveillance.”* — **IPC Focal Person**

### 3. INFRASTRUCTURE AND RESOURCE SURVEILLANCE

Interviews revealed that surveillance is not only a matter of tracking pathogens but also of monitoring the basic structural requirements for IPC. There is a perceived disconnect between identifying a resource gap and the subsequent administrative action required to fix it:

> *“We should have frequent supervision and even implementation. Because when I say we don’t have the paper towels it ends there.”* — **IPC Team Member**

### 4. HUMAN RESOURCE CONSTRAINTS AND THE RATIO OF CARE

A critical systemic barrier identified was the severe shortage of dedicated IPC personnel. Current staffing levels at Thika Level 5 Hospital deviate significantly from national and international guidelines, which recommend at least one coordinator per 250 beds:

> *“Here you find one focal person against 500 beds. This person is not just dealing with IPC alone but also their other tasks …So, it becomes hard to conduct surveillance.”* — **IPC Focal Person**

### 5. THE NEED FOR UNIT-LEVEL DECENTRALIZATION

To mitigate the challenges of staff rotation and inconsistent training, participants advocated for the permanent placement of IPC focal persons within every hospital unit. This was seen as a solution to the duty-based nature of nursing and clinical work, which often leaves rotating staff unaware of specific unit demands:

> *“We want the IPC unit in every unit. Because… today a person is there, and tomorrow another is there, and they may not have been instructed on the demands of IPC.”* — **IPC Team Member**

## BEHAVIORAL DRIVERS: ATTITUDE, IGNORANCE, AND PROFESSIONAL ACCOUNTABILITY

A significant theme emerged regarding the human element of IPC adherence. Participants noted that poor practices are not always rooted in a lack of technical knowledge, but rather in a combination of professional attitude, *“silly”* oversights, and occasional ignorance of the consequences.

### 1. THE GAP BETWEEN KNOWLEDGE AND PRACTICE

Healthcare providers observed that staff often possess the necessary training but fail to translate that knowledge into consistent action at the bedside. This suggests that attitude serves as a hidden barrier that simple training sessions may not fully address:

> *“Because when we are dealing with multitudes, sometimes you can see that person doesn’t know that he is supposed to perform hand hygiene, but you can also see attitude and ignorance.”* — **Nurse Manager, New Born Unit**

### 2. HUMAN ERROR AND NORMATIVE BEHAVIOR

Participants expressed a high degree of self-awareness regarding the fallibility of staff. They highlighted that even in high-stakes environments like the Newborn Unit, lapses in protocol are sometimes dismissed as human errors rather than technical failures:

> *“Be it a mother, be it a staff, we are human and sometimes we don’t do the wrong thing because we don’t know, but sometimes we are silly.”* — **Nurse Manager, Newborn Unit**

### 3. EXTENDING ACCOUNTABILITY TO THE COMMUNITY

Interestingly, some participants felt that addressing ignorance requires a continuum of care that extends beyond the hospital walls. They advocated for community health practitioners to reinforce these behaviors at home to ensure that IPC becomes a cultural norm rather than just a clinical requirement:

> *“But I think community health practitioners should be given the go ahead to teach them (community) even back at home. Because some of them it is not that they don’t know, it’s just ignorance.”* — **IPC Committee Member**

## DISCUSSIONS

By mapping the qualitative themes that emerged from this study, we propose a conceptual framework, Fig 1, illustrating how macro-level systemic failures trickle down to dictate frontline clinical behavior. Rather than viewing resource scarcity or poor protocol compliance as isolated behavioral deficits, this model demonstrates a distinct causal pathway: top-down policy lags and structural procurement latencies directly manifest as acute workforce strain and clinical triage at the point of care. Consequently, frontline healthcare workers are forced into compromised, improvised practices that structurally disconnect them from national safety standards. By visualizing these dynamics, the framework shifts the paradigm of IPC failure from individual professional accountability to a predictable consequence of a disabling institutional ecosystem, offering a diagnostic map for targeted, systems-level interventions.

**Fig1.**
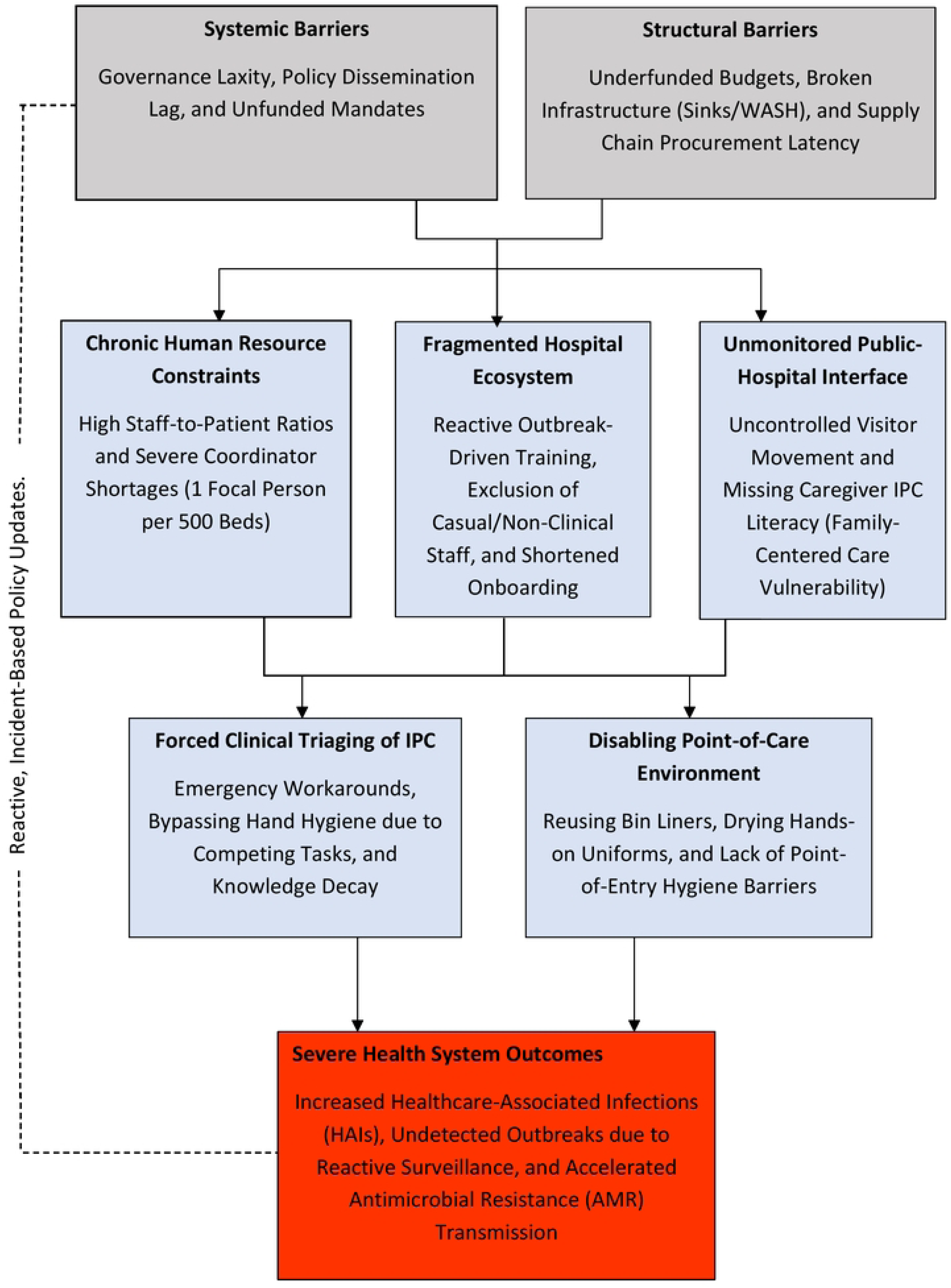
Conceptual framework of systematic and structural barriers to IPC practices. Causal pathway illustrating how macro-level policy lags and procurement latencies manifest as workforce strain, driving improvised practices at point of care

### THE IMPLEMENTATION-POLICY GAP AND STRUCTURAL GOVERNANCE

Our findings reveal a profound implementation-policy gap at facility A. While Kenya has established a comprehensive national IPC framework [5], high-level policy development frequently outpaces the financial and human resource capacities of frontline facilities.

This friction is exemplified by the local IPC committee’s inability to enforce guidelines due to a lack of institutional backing. This stands in direct opposition to the organizational framework proposed by [21], who argue that an IPC committee must serve as the primary administrative authority empowered to mandate evidence-based changes. Instead, administrative laxity and dissemination bottlenecks, such as receiving policies an entire year later, foster an environment where outdated, traditional practices persist. As [22] emphasize, institutional and administrative commitment is just as vital as technical guidelines themselves in establishing a sustainable culture of safety.

### WORKFORCE STRAIN AND REACTIVE SURVEILLANCE

A critical structural barrier identified in this study is extreme workforce strain, characterized by a staggering 1:500 IPC focal person-to-bed ratio, more than double the national recommendation [5]. This shortage forces a dangerous triaging of IPC and gives rise to the lone provider phenomenon in high-risk zones like the Newborn Unit. This severe understaffing directly cripples the facility’s capacity for proactive surveillance, reducing monitoring to a reactive mechanism triggered only by visible crises or outbreaks.

Without a reliable epidemiological baseline, hospital management cannot benchmark Healthcare-Associated Infections (HAIs) or make data-driven decisions. Nationally, [9] and [23] corroborate that these exact systemic resource gaps, supply chain bottlenecks, and severe staffing deficits are the primary drivers forcing Kenyan healthcare workers to bypass standard protocols in favor of clinical survival.

### THE DISABLING ENVIRONMENT: INFRASTRUCTURE VS. BEHAVIORAL DRIVERS

The operational reality of the hospital’s physical layout creates what participants call a disabling environment. This structural decay is highlighted by the complete failure of hand-hygiene stations, such as the female surgical ward where all six sinks were non- functional. Quantitative data from [24] confirms that a lack of functional Water, Sanitation and Hygiene (WASH) access and a shortage of drying materials remain pervasive structural barriers across Kenyan public hospitals.

Importantly, our data shows that this infrastructure failure has a cascading effect on healthcare worker behavior. While unit managers frequently cited ignorance, attitude, or shortcuts as barriers, these individual behaviors cannot be viewed in isolation. In alignment with the WHO Multimodal Strategy, behavior is heavily conditioned by system change (the physical availability of tools) and workplace enablers. When a clinician is systematically forced to reuse bin liners or dry clean hands-on contaminated clothing, a classic zero-work phenomenon, the professional safety culture erodes.

Decades of global literature, from landmark meta-analyses by [25] to regional studies by [26], [27], and [22], validate this exact paradox: even when healthcare workers maintain excellent theoretical knowledge and positive attitudes toward IPC, institutional failures and poor procurement systems inevitably override individual intentions and force a reversion to sub-optimal practices [22,25–27].

### DECENTRALIZATION AND THE EXPANDED IPC ECOSYSTEM

Finally, an emergent theme from our study underscores the need to expand the traditional boundaries of the IPC ecosystem to include the public and rotating medical students. Participants identified family caregivers as a critical population capable of vectoring infections due to unmonitored movement across overcrowded wards [28]. Furthermore, the observation that clinical students arrive with minimal IPC training points to an urgent gap in early medical and nursing curricula [29]. To bridge these gaps, IPC governance must be decentralized. Transitioning from a centralized, overburdened oversight committee to embedding dedicated IPC focal persons within every distinct ward unit would facilitate the continuous, localized sensitization required to safeguard permanent staff, rotating students, and the communities they serve.

## LIMITATIONS

This study is limited by its small sample size of seven key informants at a single facility. While this limits the generalizability of the findings, the depth of the qualitative insights provided by committee members and department managers offers a nuanced understanding of the systemic failures that quantitative surveys might miss.

## RECOMMENDATIONS

To bridge the gap between national IPC policies and facility-level compliance, health systems must transition from a centralized oversight model to a decentralized, ward-level governance structure. Appointing dedicated *IPC Champions*, (such as staff nurses, clinical officers, or department leads embedded within specific units), provides a mechanism for real-time, peer-to-peer supervision. In resource-constrained settings where central IPC committees are frequently understaffed and structurally detached from daily ward dynamics, these champions serve as immediate focal points. They can monitor hand hygiene compliance, audit sterile processing workflows, and troubleshoot supply stockouts (such as PPE or alcohol-based hand rubs) at the exact point of care. Furthermore, by framing IPC enforcement as a shared, localized responsibility rather than a top-down bureaucratic mandate, this approach fosters an internal culture of patient safety and immediate accountability.

Transitioning to digital health platforms bridges the policy-to-practice gap by instantly delivering updated guidelines to frontline clinicians, replacing outdated paper protocols with real-time mobile alerts and electronic records. This digital modernization ensures immediate, facility-wide alignment with evolving national infection control standards.

Simultaneously, establishing formal IPC briefings for patient relatives addresses a critical vulnerability in health systems heavily reliant on family-centered bedside care. In settings where family members routinely provide essential, hands-on patient support, basic infrastructure limitations can amplify transmission risks. Targeted, culturally accessible education on hand hygiene and isolation protocols mitigate these structural gaps. Ultimately, integrating digital dissemination for healthcare staff with robust community engagement for visitors transforms informal care networks from potential vectors of cross-contamination into active, informed partners in preventing healthcare-associated infections and curb antimicrobial resistance at the point of care.

## CONCLUSION

This study demonstrates that while Kenya possesses a robust national infection prevention and control (IPC) framework, a profound policy-to-practice lag persists at the institutional level. This implementation gap is driven by interconnected systemic and structural barriers rather than a singular clinical failure. Chronic understaffing forcing a critical triaging of IPC measures, prolonged procurement latency causing severe infrastructure decay, and a reactive, sporadic training culture collectively create a disabling environment for frontline healthcare workers. Furthermore, the study highlights critical vulnerabilities at the community-hospital interface, where unmonitored, family-centered care networks inadvertently function as vectors for cross-contamination due to a lack of structured public sensitization.

To bridge this gap and reduce the substantial burden of healthcare-associated infections (HAIs), hospital leadership and county health departments must transition from top-down, crisis-driven mandates to localized, proactive structural enablers. First, IPC oversight must be decentralized through the appointment of ward-level champions to facilitate continuous peer-supervision and address resource deficits in real time.

Second, the traditional, workshop-reliant training model must be replaced with sustainable, low-cost digital health integrations, such as mobile updates or clinical networks, to prevent knowledge decay and ensure rapid protocol dissemination. Third, targeted, culturally accessible IPC education must be institutionalized for informal caregivers to secure the patient-visitor environment. Finally, continuous surveillance must be properly funded and staffed to shift institutional behavior from reactive outbreak management to data-driven, proactive prevention. Ultimately, national IPC strategies cannot remain rigid, idealized mandates; they must be dynamically adapted, funded, and tailored to the nuanced, everyday operational realities and structural limitations faced by clinical staff within low-resource referral facilities.

## Data Availability

All relevant data underlying the findings of this study are fully contained within the manuscript and its supporting information file.

## ACKNOWLEDGEMENTS

We would like to express our sincere gratitude to the hospital administration and departmental health managers at facility A for granting permission and providing facilities to conduct this research. We are deeply indebted to the healthcare providers and IPC committee members who generously shared their time, lived experiences, and invaluable insights during the key informant interviews. We also thank the faculty and research leadership at the United States International University-Africa (USIU-Africa) for their guidance and institutional support throughout the conceptualization and execution of this study.

## Supporting Information

S1 File. Qualitative Thematic Framework & Clinical Implications

## Notes

### Competing Interest Statement

The author have the following compteting interests to declare: Miss Janet Ngethe, Nicole Wamaitha, and Cynthia Nduta serve as advisors for the Ministry of Health, Kiambu County Governemnt in relation to infection prevetntion and control policy development. The remaing authors declare no competing interests exist.

### Author Declarations

Ethical approval was received from all these institutions: 1. Institutional Review Board (IRB) ethics clearance certificate from the United States International University- Africa 2. National Commission for Science, Technology and Innovation research license from The Government of Kenya. 3. Kiambu Health Department license from the Kiambu County Government 4. Thika Level 5 Teaching and Referral Hospital approval letter, from Thika level 5 hospital.

## REFERENCES

1. World Health Organization. Guidelines for the prevention and control of carbapenem-resistant Enterobacteriaceae, Acinetobacter baumannii and Pseudomonas aeruginosa in health care facilities. Geneva: World Health Organization; 2017. Available from: https://www.who.int/publications/i/item/guidelines-for-the-prevention-and-control-of-carbapenem-resistant-enterobacteriaceae-acinetobacter-baumannii-and-pseudomonas-aeruginosa-in-health-care-facilities

2. World Health Organization. Global action plan and monitoring framework on infection prevention and control (IPC) 2024–2030. Geneva: World Health Organization; 2024. Available from: https://www.who.int/teams/integrated-health-services/infection-prevention-control/draft-global-action-plan-and-monitoring-framework-on-ipc

3. World Health Organization. Global report on infection prevention and control 2024. Geneva: World Health Organization; 2024. Available from: https://www.who.int/publications/i/item/9789240051164

4. Melariri H, Freercks R, van der Merwe E, Ham-Baloyi WT, Oyedele O, Murphy RA, et al. The burden of hospital-acquired infections (HAI) in sub-Saharan Africa: a systematic review and meta-analysis. EClinicalMedicine. 2024;71:102571. 10.1016/j.eclinm.2024.102571

5. Ministry of Health. Kenya national infection prevention and control strategic plan for health care services 2021–2025. 2nd ed. Nairobi: Government of Kenya; 2021. Available from: http://guidelines.health.go.ke:8000/media/Kenya_National_IPC_Strategic_Plan_for_Health_Care_Services_2021_-_2025.pdf

6. University of Nairobi, Department of Paediatrics & Child Health. Infection prevention and control (IPC). Nairobi: University of Nairobi; 2021. Available from: https://paediatrics.uonbi.ac.ke/sites/paediatrics.uonbi.ac.ke/files/2023-02/2.%20IPC%20-%20April%202021.pdf

7. World Health Organization, United Nations Children’s Fund (UNICEF). Water, sanitation, hygiene, and waste management for SARS-CoV-2, the virus that causes COVID-19: Interim guidance. 3rd ed. Geneva: World Health Organization; 2020. Available from: https://iris.who.int/server/api/core/bitstreams/6cfcc886-53dc-4168-9669-d6617ee1b443/content

8. World Health Organization. WHO calls for better hand hygiene and other infection control practices. Geneva: World Health Organization; 2021. Available from: https://www.who.int/news/item/05-05-2021-who-calls-for-better-hand-hygiene-and-other-infection-control-practices

9. Brown H, Odhiambo A, Mwaki A, Atieno N, Ouda R, Ngere I. Understanding the drivers of poor infection prevention control (IPC) practices in Kenyan health facilities: An interdisciplinary study. PLOS Global Public Health. 2025;5(6):e0004404. 10.1371/journal.pgph.0004404

10. Ministry of Health Kenya. National infection prevention and control policy for health care services in Kenya. Nairobi: Government of Kenya; 2010. Available from: http://guidelines.health.go.ke:8000/media/infection_control_policy.pdf

11. Ministry of Health. Interim guidelines on management of COVID-19 in Kenya. Nairobi: Government of Kenya; 2020. Available from: https://www.health.go.ke/wp-content/uploads/2020/06/Updated-Case-Management-Guidelines-26_03_20-1.pdf

12. World Health Organization. Guidelines on core components of infection prevention and control programmes at the national and acute health care facility level. Geneva: World Health Organization; 2016. Available from: https://iris.who.int/server/api/core/bitstreams/a4246b85-af71-4b03-98c1-7a2b6c850029/content

13. Lowe H, Woodd S, Lange IL, Janjanin S, Barnet J, Graham W. Challenges and opportunities for infection prevention and control in hospitals in conflict-affected settings: A qualitative study. Conflict and Health. 2021;15(1):94. 10.1186/s13031-021-00428-8

14. Arora V, Singh B, Sangam K, Hasmee N, Gurung M. Overcoming barriers to infection prevention and control compliance in intensive care units: A call for strategic change. Nursing in Critical Care. 2025;30(3):e70012. 10.1111/nicc.70012

15. Weiss J, Berwa S, Momanyi G, Nduwayezu R, Siraj D, Shirley D. Barriers and facilitators to infection prevention and control practices at King Faisal Hospital, Kigali, Rwanda. Antimicrobial Stewardship & Healthcare Epidemiology. 2025;5(1):e197. 10.1017/ash.2025.10111

16. Abbas S. The challenges of implementing infection prevention and antimicrobial stewardship programs in resource-constrained settings. Antimicrobial Stewardship & Healthcare Epidemiology. 2024;4(1):e35. 10.1017/ash.2024.35

17. World Health Organization. Global report on infection prevention and control. Geneva: World Health Organization; 2022. Available from: https://www.who.int/publications/i/item/9789240051164

18. World Health Organization. Improving infection prevention and control at the health facility: Interim practical manual supporting implementation of the WHO Guidelines on Core Components of Infection Prevention and Control Programmes. Geneva: World Health Organization; 2018.

19. Nyaga A. Adherence to infection prevention and control guidelines among nurses in Thika level 5 hospital, Kiambu county, Kenya [Thesis]. Thika: Mount Kenya University; 2023. Available from: https://erepository.mku.ac.ke/handle/123456789/7031

20. Office of the Auditor-General Kenya. Summary report of the Auditor-General on Level 4 and Level 5 hospitals in Kenya: Financial year 2021/2022. Nairobi: Office of the Auditor-General; 2024. Available from: https://www.oagkenya.go.ke/wp-content/uploads/2024/03/Summary-Book-for-Level-4-and-Level-5-Hospitals.pdf

21. Kubde D, Badge AK, Ugemuge S, Shahu S. Importance of hospital infection control. Cureus. 2023;15(12):e50931. 10.7759/cureus.50931

22. Ider BE, Clements A, Adams JS, Whitby M, Muugoltsog O. Perceptions of healthcare professionals regarding the main challenges and barriers to effective hospital infection control in Mongolia: a qualitative study. BMC Infectious Diseases. 2010;10(1):1–10. 10.1186/1471-2334-10-244

23. Asgedom AA, Ruano AL, Moen BE. Assessment of infection prevention and control in public hospitals in the post war Tigray region of Ethiopia using the WHO Infection Prevention and Control Assessment Framework (IPCAF). Antimicrobial Resistance and Infection Control. 2026;15(1):82. 10.1186/s13756-026-01746-3

24. Maina M, Tosas-Auguet O, McKnight J, Zosi M, Kimemia G, Mwaniki P, et al. Extending the use of the World Health Organisations’ water sanitation and hygiene assessment tool for surveys in hospitals - from WASH-FIT to WASH- FAST. PLOS One. 2019;14(12):e0226548. 10.1371/journal.pone.0226548

25. Allegranzi B, Nejad SB, Combescure C, Iten A, Kilpatrick C, Cookson B, et al. Burden of endemic health-care-associated infection in developing countries: systematic review and meta-analysis. The Lancet. 2011;377(9761):228–241. 10.1016/S0140-6736(10)61458-4

26. Kindu M, Yenesew MA, Kassie GG. Infection prevention practices and associated factors among healthcare professionals in West Gojjam Zone public Hospitals Northwest Ethiopia, 2023. PLOS One. 2026;21(1):e0338621. 10.1371/journal.pone.0338621

27. Iliyasu G, Dayyab FM, Habib ZG, Tiamiyu AB, Abubakar S, Mijinyawa MS, et al. Knowledge and practices of infection control among healthcare workers in a Tertiary Referral Center in North-Western Nigeria. Annals of African Medicine. 2016;15(1):34–40. 10.4103/1596-3519.161724

28. Marquer C, Guindo O, Mahamadou I, Job E, Rattigan SM, Langendorf C, et al. An exploratory qualitative study of caregivers’ knowledge, perceptions and practices related to hospital hygiene in rural Niger. Infection Prevention in Practice. 2021;3(3):100160. 10.1016/j.infpip.2021.100160

29. Ojulong J, Mitonga KH, Iipinge SN. Knowledge and attitudes of infection prevention and control among health sciences students at University of Namibia. African Health Sciences. 2014;13(4):1071–1078. 10.4314/ahs.v13i4.30

